# Integrative Metabolomic and Proteomic Signatures Define Clinical Outcomes in Severe COVID-19

**DOI:** 10.1101/2021.07.19.21260776

**Authors:** Mustafa Buyukozkan, Sergio Alvarez-Mulett, Alexandra C. Racanelli, Frank Schmidt, Richa Batra, Katherine L. Hoffman, Hina Sarwath, Rudolf Engelke, Luis Gomez-Escobar, Will Simmons, Elisa Benedetti, Kelsey Chetnik, Guoan Zhang, Edward Schenck, Karsten Suhre, Justin J. Choi, Zhen Zhao, Sabrina Racine-Brzostek, He S. Yang, Mary E. Choi, Augustine M.K. Choi, Soo Jung Cho, Jan Krumsiek

## Abstract

The novel coronavirus disease-19 (COVID-19) pandemic caused by SARS-CoV-2 has ravaged global healthcare with previously unseen levels of morbidity and mortality. To date, methods to predict the clinical course, which ranges from the asymptomatic carrier to the critically ill patient in devastating multi-system organ failure, have yet to be identified. In this study, we performed large-scale integrative multi-omics analyses of serum obtained from COVID-19 patients with the goal of uncovering novel pathogenic complexities of this disease and identifying molecular signatures that predict clinical outcomes. We assembled a novel network of protein-metabolite interactions in COVID-19 patients through targeted metabolomic and proteomic profiling of serum samples in 330 COVID-19 patients compared to 97 non-COVID, hospitalized controls. Our network identified distinct protein-metabolite cross talk related to immune modulation, energy and nucleotide metabolism, vascular homeostasis, and collagen catabolism. Additionally, our data linked multiple proteins and metabolites to clinical indices associated with long-term mortality and morbidity, such as acute kidney injury. Finally, we developed a novel composite outcome measure for COVID-19 disease severity and created a clinical prediction model based on the metabolomics data. The model predicts severe disease with a concordance index of around 0.69, and furthermore shows high predictive power of 0.83-0.93 in two previously published, independent datasets.

## Introduction

The novel coronavirus disease 2019 (COVID-19) has a broad spectrum of clinical features that range from asymptomatic disease to acute respiratory distress syndrome (ARDS)(1, 2). COVID-19 ARDS can lead to refractory hypoxia, mechanical ventilation, prolonged intensive care unit (ICU) stay and increased mortality(3). Previous studies have shown a high incidence of concomitant organ failure in COVID-19, including acute kidney injury (AKI)(4), acute liver injury(5), thromboembolic events(6, 7) and secondary infections contributing to a fatal outcome(8).

Massive investigative efforts by multiple scientific groups have used proteomic and metabolomic approaches to begin to unravel disease mechanisms relevant to SARS-CoV-2 infection such as inflammation, coagulation, and metabolism(9). However, how COVID-19 specific protein-metabolite interactions relate to the severity of disease and clinical outcomes remains poorly understood. Key study limitations have included relatively small sample sizes, absence of protein-metabolite network analysis and the focus on dichotomous outcome measures such as death and survival. These limitations have been difficult to overcome and restrict our understanding of COVID-19 pathogenesis.

Here, we report the largest study to integrate targeted metabolomic and proteomic analyses of serum samples obtained from hospitalized COVID-19 patients during SARS-CoV-2 infection compared to patients admitted during the same time period with symptoms related to COVID-19 and negative RT-PCR for SARS-CoV-2 as controls (Figure 1). Through this work, we uncovered COVID-19-specific metabolite and protein profiles, identified novel protein-metabolite modules, and defined the molecular signatures of several clinical indices (CRP, ferritin, platelet count, AKI, and death). Additionally, we developed the first clinical composite outcome prediction model in COVID-19, where the input of discrete metabolic profiles significantly improves our clinical insight into a broad range of outcomes known to plague many survivors of severe COVID-19.

**Fig. 1.**
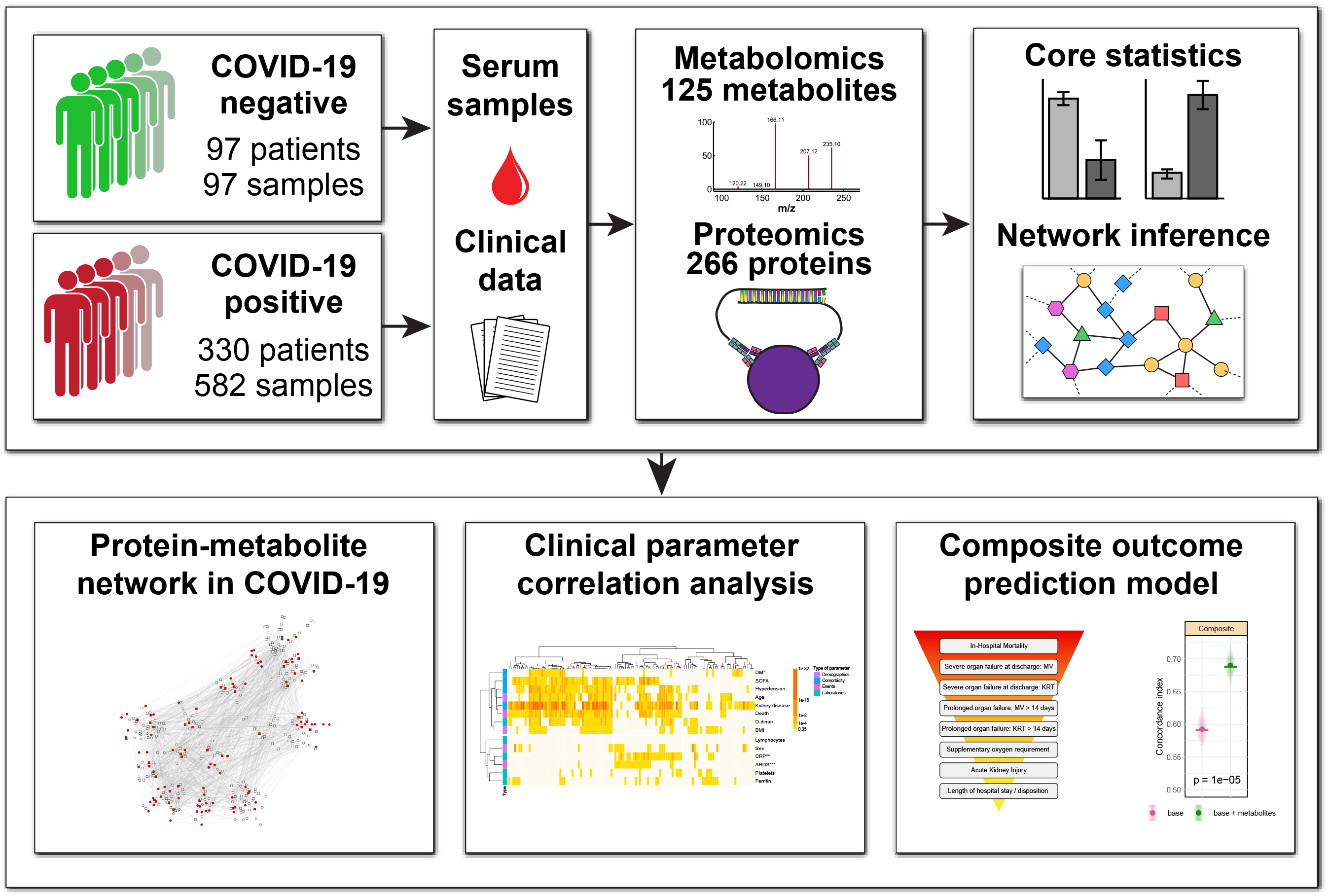
Study outline. Study population composed by COVID-19 patients and controls. Serum samples obtained from clinically indicated specimens during the first 72 hours of admission. Metadata obtained from electronic medical records. We obtained metabolomic and proteomic profile, after which we integrated our findings into a single network describing the associations between differentially expressed proteins and metabolites in COVID-19. We then correlated the metabolites and proteins with baseline characteristics, laboratory parameters and clinical events. Finally, we developed a novel metabolite-based prediction model for a composite outcome measure comprised of key clinical parameters including death, mechanical ventilation, initiation of dialysis, supplemental oxygen requirement, development of acute kidney injury, and length of hospital stay.

## Results

### Study cohort and dataset

Our cohort was comprised of 330 patients with confirmed SARS-CoV-2 RT-PCR, and 97 non-COVID-19 controls with negative RT-PCR results who were hospitalized at the NewYork-Presbyterian Hospital/Weill Cornell Medical Center between March and April 2020. Serum samples were obtained within the first 3 days of admission. The majority of COVID-19 patients had samples drawn at two or three different time points resulting in a total of 582 serum samples from the 330 COVID-19 patients, while all 97 controls had one sample drawn. Metabolomics was measured for all available samples. Proteomics was measured for fewer samples (n=189), also across different time points for some patients. Notably, there were only minor time effects across the three days (Supplementary Figure 1), and the repeated samples were thus treated as replicates using a linear mixed effect model, see Methods. A detailed description of the clinical and demographic characteristics of the cohort can be found in Table 1, Supplementary Table 1 and Supplementary Table 2. Of note, we excluded samples collected after intubation because we found that the clinical act of intubation significantly alters a patient’s metabolic profile (Supplementary Table 3).

Metabolic profiles were assessed for all samples using liquid chromatography coupled with mass spectrometry (LC/MS). After quality control and data preprocessing, 125 metabolites were available for comparative analysis. Targeted proteomic profiling was performed on a subset of 227 samples (173 from COVID-19 patients and 54 controls) using the Olink inflammation, cardiovascular II and cardiovascular III panels, which cover 266 unique protein biomarkers. These panels were selected since it has previously been shown that inflammation and cardiovascular pathways are essential during COVID-19 pathogenesis(10).

### Metabolomic and proteomic changes associated with COVID-19

Differential metabolomic analysis identified significant changes in abundance of 70 out of the 125 analyzed metabolites between COVID-19 and controls at a false discovery rate (FDR) of 0.05 (Figure 2A). The top three differentially expressed metabolites were involved in amino acid metabolism: N-acetyl-L-aspartic acid (p-value = 9.19E-18), N-acetyl-aspartyl-glutamic acid (p-value = 5.30E-15), and argininosuccinic acid (p-value = 6.35E-12) (Figure 2B). KEGG pathway mapping of the differentially expressed metabolites revealed an involvement of various metabolic pathways (Figure 2C). These pathways included arginine and proline metabolism, glycine and serine metabolism, alanine metabolism, methionine metabolism, sphingolipid metabolism, gluconeogenesis, and the TCA cycle pathway, demonstrating involvement of the broader categories of amino acid, lipid and energy metabolism in COVID-19 pathogenesis. Our results confirm prior reports that reported altered phenylalanine and tryptophan metabolism in severe COVID-19 patients compared to non-COVID-19 patients(11-13). Additionally, our data has shown that multiple metabolites involved in sphingolipid metabolism are significantly increased in COVID-19 patients, pointing towards other potential targets for COVID-19 treatment(14).

**Fig. 2.**
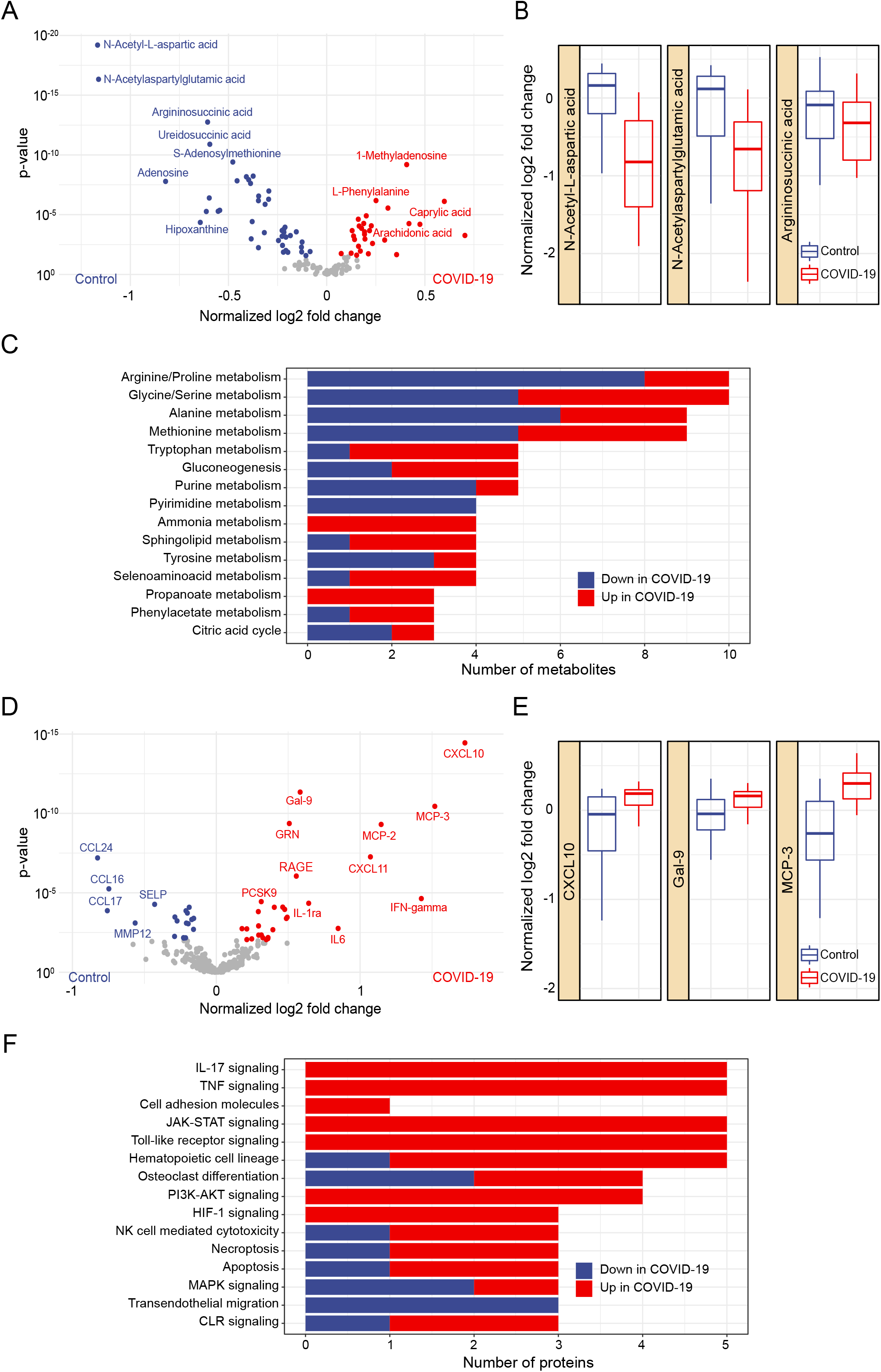
Metabolomics and Proteomics changes associated with COVID-19. **A**, Volcano plot showing differentially expressed metabolites between COVID-19 patients and controls at an adjusted p-value <0.05. In red, upregulated metabolites in COVID-19 patients. In blue, upregulated metabolites in the control group. **B**, Top 3 differentially expressed metabolites in COVID-19 patients vs. controls based on adjusted p-values. Y axis shows log_2_ fold changes in relation to the mean of the control group. **C**, Number of significantly regulated molecules in KEGG pathways, top 15 shown. **D**, Volcano plot showing the differentially expressed proteins between COVID-19 patients and controls at an adjusted p-value <0.05. **E**, Top 3 differentially expressed proteins in COVID-19 patients vs. controls based on adjusted p-values. Y axis shows log_2_ fold changes in relation to the mean of the control group. **F**, Number of significantly regulated molecules in KEGG pathways, top 15 shown.

Comparative proteomic analysis identified significant changes in the expression of 48 out of the 266 analyzed proteins between COVID-19 and controls at an FDR of 0.05 (Figure 2D). The top three differentially expressed proteins were C-X-C motif chemokine ligand 10 (CXCL10) (p-value = 5.09E-13), galectin 9 (Gal-9) (p-value = 5.49E-10), and monocyte chemoattractant protein 3 (MCP-3) (p-value = 2.85E-09) (Figure 2E). KEGG pathway mapping revealed that these proteins participated in various protein pathways (Figure 2F), including the interleukin 17 (IL-17), tumoral necrosis factor (TNF) and JAK-STAT signaling pathways. Detailed results of the differential analysis and pathway mappings can be found in Supplementary Table 4. Of note, the utility of the JAK inhibitors baricitinib or ruxolitinib has already been demonstrated in a clinical trial for selected patients with severe or critical COVID-19 patients(15, 16). Based on our data, clinical trials further targeting IL-17 and TNF signaling in COVID-19 may lead to additional therapeutic approaches for treating COVID-19.

Global principal component analysis (PCA) on the metabolomics and proteomics data revealed no clear separation of COVID-19 and control groups (Supplementary Figure 2). This is an effect that we have commonly observed in previous studies of blood data(17-22), where omics profiles only separated groups in a specific (single molecules and pathways) rather than a global fashion.

### Protein-metabolite networks identify potential mediators of COVID-19 pathology

To obtain further mechanistic insight into the biology of COVID-19, we developed a comprehensive, data-driven network for the integrative analysis of our multi-omics dataset. We first generated a Gaussian graphical model (GGM) of correlated metabolites and proteins from our COVID-19 cohort (Supplementary Data 1). GGMs are correlation-based network models that we have previously demonstrated to accurately reconstruct biological pathways from blood-based omics data(23-25). A minimum spanning tree-based algorithm was then used to identify a focused subnetwork that connects the most significantly correlated metabolites and proteins from the original network (Figure 3). The subnetwork included 13 proteins from the Olink inflammatory panel, 32 proteins from the Olink cardiovascular panels II and III, and 70 metabolites. From this subnetwork, we selected 4 network modules to query the interplay between metabolism, inflammation, and vascular dysfunction in COVID-19 patients.

**Fig. 3.**
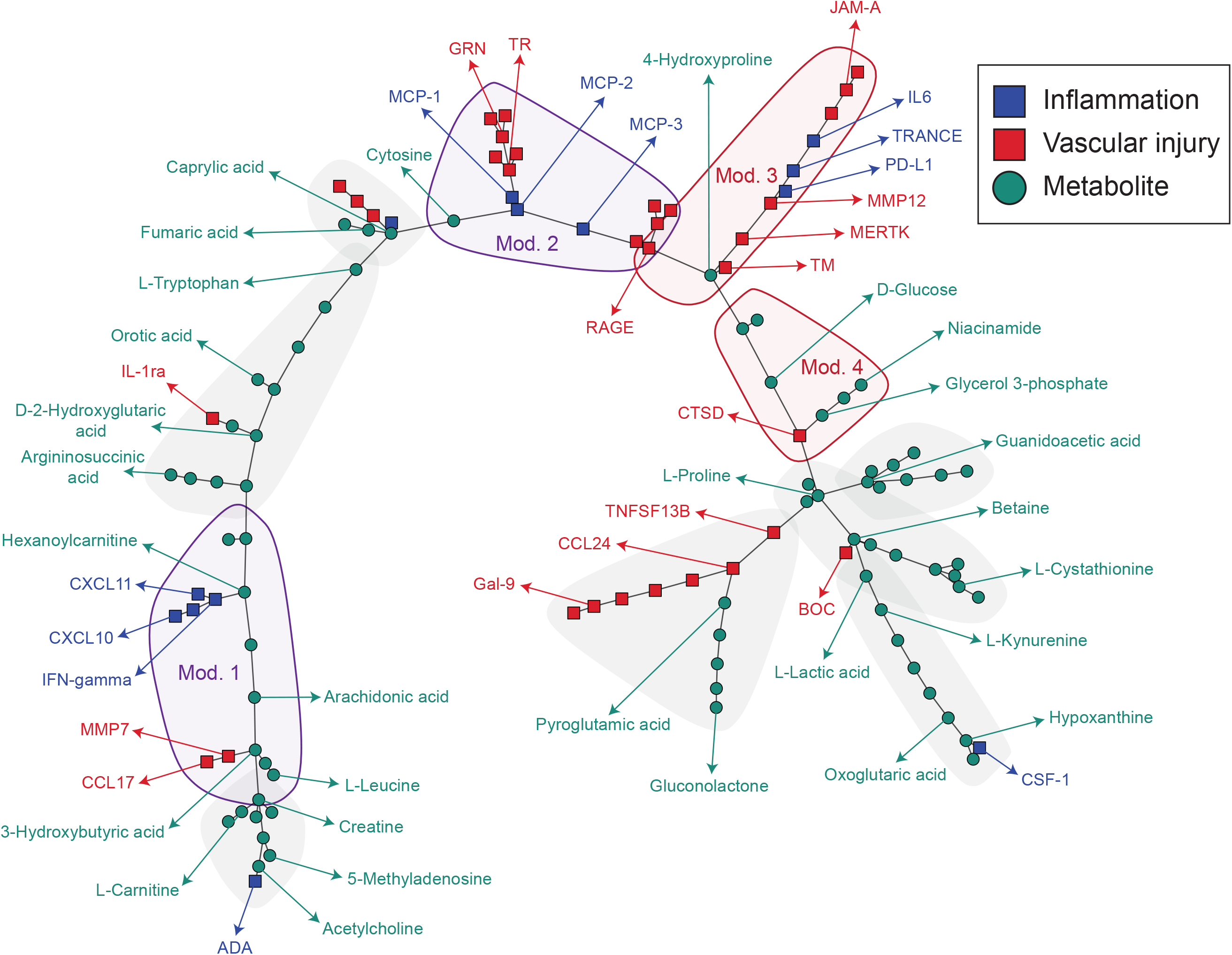
Protein-metabolite networks in COVID-19. Gaussian graphical model (GGM) representing the significant partial correlations between all the measured metabolites and proteins, generated by a minimum spanning tree (MST) based reduction combined with a shortest paths-based approach. The network includes all differentially expressed metabolites and proteins between COVID-19 patients and controls. Squares indicate proteins and green circles indicate metabolites. Inflammatory proteins are colored in purple, while vascular injury proteins are colored in red. Shadows represent molecule modules.

#### Inflammation-related network modules Module 1

Our network identified hexanoylcarnitine as a key metabolite associated with inflammatory cytokines in COVID-19 illness. The module contained IFN-gamma, CXCL10 and CXCL11 that were all upregulated in COVID-19 and associated with hexanoylcarnitine. IFN-gamma, CXCL10 and CXCL11 are proinflammatory cytokines which regulate T cell immunity(26), while hexanoylcarnitine is a medium-chain fatty acid conjugate that plays a critical role in energy metabolism and mitochondrial fatty acid β-oxidation (Figure 3)(27). Levels of other carnitine species (L-carnitine and caprylic acid) were also elevated in COVID-19 patients, suggesting a role for inflammation, dysregulated fatty acid β-oxidation and mitochondrial dysfunction in COVID-19 pathogenesis(12, 28-31).

#### Module 2

In a second inflammation-related module, cytosine was another key metabolite linked with inflammatory cytokines during COVID-19. The module consisted of a group of macrophage-derived cytokines, including MCP-2, MCP-3 and GRN, which were all upregulated in COVID-19 and positively associated with cytosine (Figure 3). This finding is consistent with the hyperinflammatory state of COVID-19 infection in which cytokine storm is often observed(32). Cytosine, a pyrimidine-class nucleotide that is an essential metabolite for cell proliferation and survival, is commonly upregulated in the host response during viral infection and is furthermore an important mediator of viral replication(33). Prior reports have shown cytosine levels to be elevated in COVID-19 patients(34), which was corroborated in our own cohort. Taken together, cytosine may be a key metabolite linking viral replication to SARS-CoV-2 induced inflammation.

#### Vascular-related network modules. Module 3

While COVID-19 presents itself mainly as a respiratory disease, autopsy reports have additionally described significant vascular injury due to endothelial cell damage, microcirculatory thrombi, and impaired cellular junction integrity(35). Our network uncovered the coordination of MERTK, RAGE and thrombomodulin (TM) with the metabolite 4-hydroxyproline in COVID-19 (Figure 3). MERTK, RAGE and TM are proteins key to maintaining features of endothelial homeostasis, and hydroxyproline is a major component of collagen(36). MERTK, a member of the TAM family of receptor tyrosine kinases (RTKs), and RAGE, a pro-coagulant and inflammatory molecule were upregulated, while TM was downregulated, suggesting induction of a pro-coagulant and inflammatory vascular state in COVID-19. Notably, the coordination of these proteins centered around alterations in levels of hydroxyproline, where downregulation of TM directly correlated with hydroxyproline levels. A link between TM and hydroxyproline has been described, where administration of TM had anti-fibrotic effects on lung and kidney murine models(37). Taken together, these data link the pro-coagulant and fibrotic state of COVID-19 through thrombomodulin and hydroxyproline.

#### Module 4

In a second vascular-related module, we found that cathepsin D (CTSD), a lysosomal protease known to disrupt endothelial cell junctions and increase vasopermeability, was associated with a group of glycolytic metabolites including D-glucose, glycerol-3-phosphate, and lactic acid(38). Notably, increased vascular permeability and glycolysis are known features of a pro-angiogenic state(39). While dysregulated angiogenesis occurs in COVID-19 patients(40), how these events are coordinated remains poorly understood. Here, we report the concomitant up-regulation of cathepsin D and intermediate products of glycolysis (D-glucose and lactic acid) in our COVID-19 cohort, which position this enzyme as a potential driver of the COVID-19 phenotype. Of note, elevated plasma activity of cathepsin D has been found in patients with type 2 diabetes, suggested a link between abnormal vasculature and the dysregulated glucose metabolism seen in our higher risk diabetic COVID-19 patients(41). Taken together these data suggest that the disruption of vascular and glucometabolic homeostasis in COVID-19 is mediated by cathepsin D.

### Serum metabolites and proteins associate with clinical indices in COVID-19

Serum metabolites and proteins within the COVID-19 patient group were assessed for correlation with relevant clinical indices including: i) demographics (sex, age, BMI), ii) concurrent comorbidity (hypertension, pre-existing kidney disease, diabetes mellitus [DM], severity of illness [SOFA]), iii) laboratory markers of inflammation (C-reactive protein [CRP], d-dimer, lymphocyte count, platelet count, ferritin), and iv) future clinical events (ARDS, death) (Figure 4A and 4D). Pre-existing kidney disease was found to associate with the highest number of both metabolites and proteins. Other clinical parameters including ARDS, death, BMI, gender, age, SOFA score, hypertension, DM, and d-dimer levels were associated with numerous metabolites but only few proteins. Conversely, platelet count and ferritin level were associated with a large number of proteins but relatively few metabolites. Detailed association results are provided in Supplementary Tables 5 and 6.

**Fig. 4.**
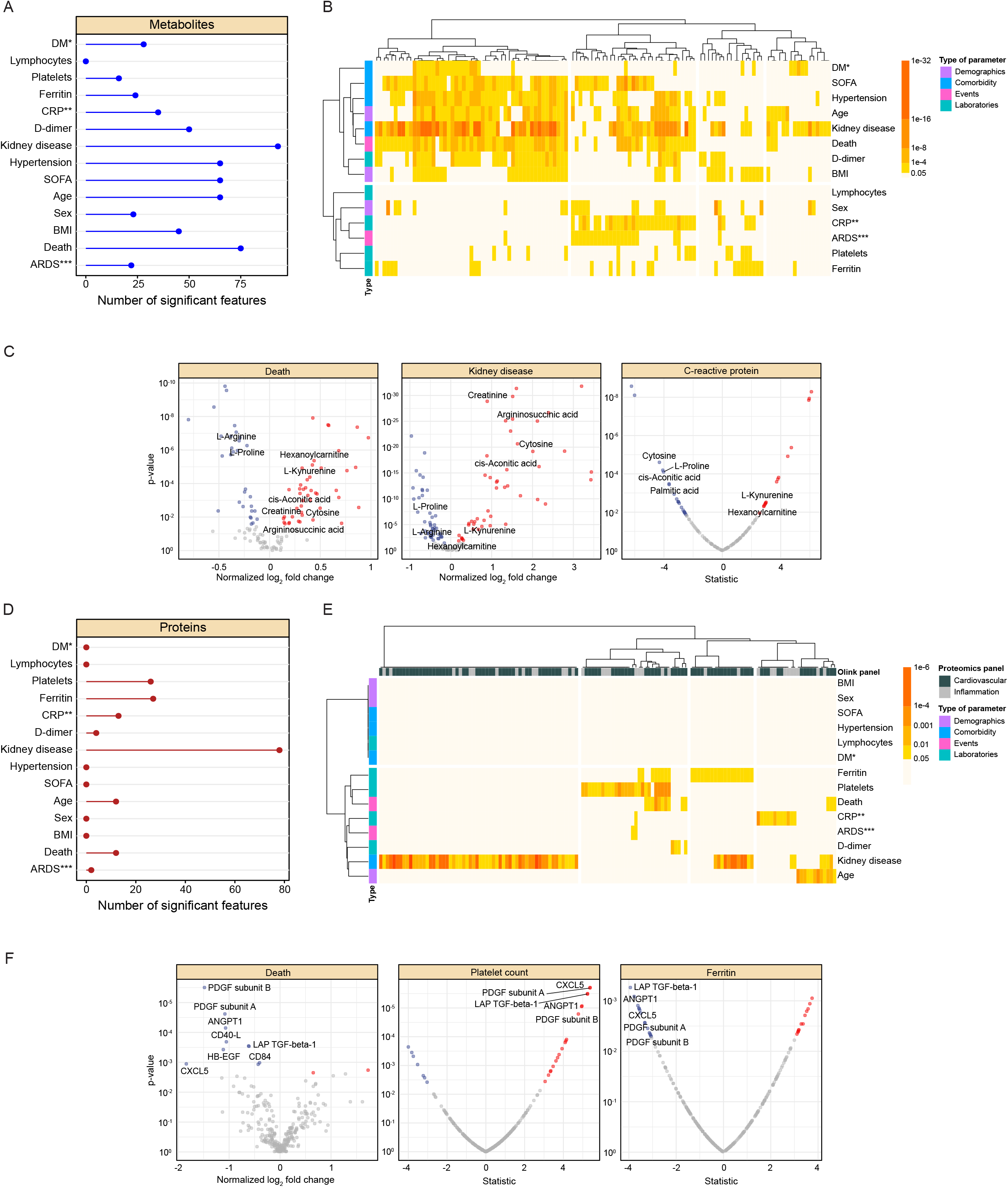
Associations of molecules with clinical indices in COVID-19. **A**, Lollipop plot representing the number of altered metabolites for each analyzed parameter (*Diabetes mellitus, **C-reactive protein, ***Acute respiratory distress syndrome, represented by whether patients were intubated). **B**, Heatmap indicating the number of differentially expressed metabolites associated with each clinical parameter and its significance. We included 4 clinical parameter categories: demographics, comorbidities, clinical events, and laboratory parameters. **C**, Volcano plot showing detailed metabolic changes correlated to death, kidney disease and C-reactive protein. **D**, Lollipop plot representing the number of altered proteins for each analyzed parameter. **E**, Heatmap indicating the number of differentially expressed proteins on each clinical parameter and its significance, using the same 4 categories described above. Proteins are marked according to Olink panels. F, Volcano plot showing detailed protein changes correlated to death, platelet count and ferritin levels.

Hierarchical clustering of clinical indices by their correlation with metabolites revealed two broad clusters: one predominantly related to laboratory markers of inflammation, and a second largely related to non-laboratory indices (Figure 4B). Volcano plots of three representative indices (death, pre-existing kidney disease and CRP) are shown in Figure 4C. Seventy-five metabolites were significantly associated with death, 96 with pre-existing kidney disease and 37 with CRP level. 69 metabolites were associated with both death and kidney disease and 20 were associated with death, kidney disease and CRP levels. Key metabolites associated with each of the three selected clinical indices are depicted in Figure 4C. Remarkably, hexanoylcarnitine and cytosine, which we earlier showed to be upregulated in COVID-19, were among the 20 metabolites associated with all three of these clinical indices. This finding supports the potential utility of hexanoylcarnitine and cytosine not only as biomarkers for COVID-19 but also as predictors of disease severity.

Clustering of clinical indices according to their correlation with proteins also revealed two broad clusters (Figure 4E) that were notably similar to the metabolite-derived clusters, except that age and kidney disease clustered together with laboratory markers of inflammation. Volcano plots of three representative indices (death, ferritin level and platelet count) are depicted in Figure 4F. Seventeen proteins correlated with death, 26 with platelet count and 29 with ferritin. Six proteins were associated with both death and ferritin level, 9 with death and platelet count, and 5 (ANGPT1, PDGF subunit A, PDGF subunit B, LAP TGF-beta-1, CXCL5) with death, ferritin level and platelet count (Figure 4F). ANGPT1, PDGF subunit A and PDGF subunit B are markers of endothelial injury and platelet dysfunction while LAP, TGF-beta-1 and CXCL5 are markers of inflammation. The fact that these proteins correlated with known clinical indices of COVID-19 mortality and morbidity corroborates the importance of vascular injury and inflammation in COVID-19 pathogenesis, which we observed in our network analysis above.

### Metabolomics signature predicts clinical outcomes in COVID-19

We devised a novel hierarchical composite outcome measure for COVID-19 severity which incorporates a series of clinical events, ranked in order of severity, that characterize both acute COVID-19 and some of the sequelae of COVID-19 seen in post-acute COVID-19 syndrome (PACS)(42): in-hospital mortality, mechanical ventilation (MV) at discharge, kidney replacement therapy (KRT) at discharge, prolonged organ failure support (mechanical ventilation and/or kidney replacement therapy for more than 2 weeks), supplemental oxygen requirement, acute kidney injury and length of hospital stay (Figure 5A,). Our measure represents each patient on a continuous spectrum of severity to provide more information than a dichotomous classification such as mortality. Details on the design of this score and patient numbers in each group can be found in Supplementary Figure 3.

**Fig. 5.**
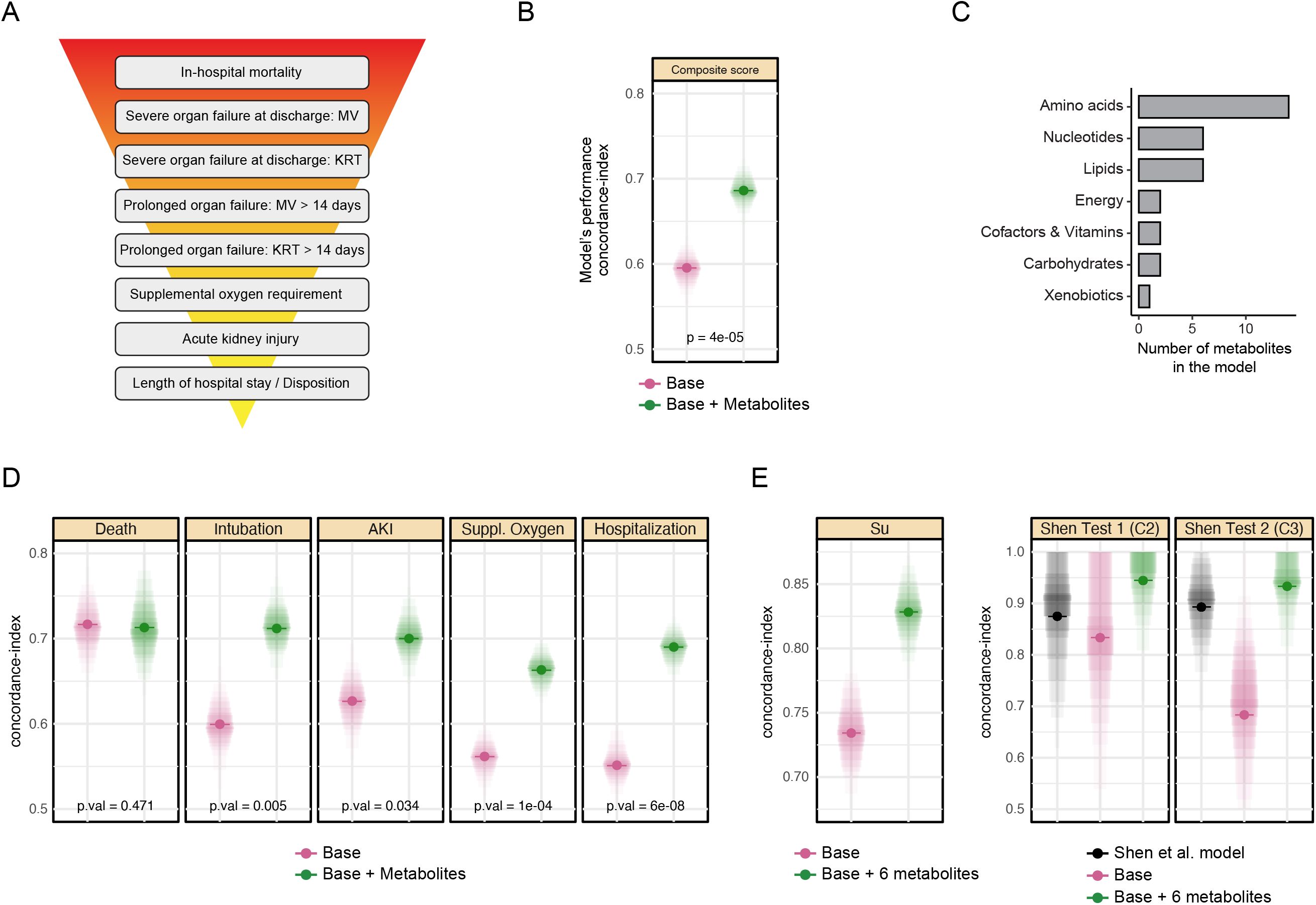
Metabolomics signature predicts clinical outcomes of COVID-19. **A**, Scheme of the hierarchical composite outcome ranging from in-hospital mortality to length of hospital stay and disposition (MV: Mechanical ventilation, KRT: Kidney replacement therapy). **B**, Dot plot comparing the predictive performance (Concordance index) of a baseline model using age, sex, and BMI and a model including baseline plus metabolites. The results show that the prediction accuracy improves when adding metabolites to the model. **C**, Bar plot showing the main altered metabolic groups included in the composite outcome score. **D**, Dot plots showing individual outcome analysis, comparing the baseline model and baseline plus metabolites added. Metabolites improve the prediction accuracy of ARDS, AKI, supplemental oxygen requirement and prolonged hospitalization. **E**, Replication analysis of our model in two independent datasets by Su et al.(43) and Shen et al.(44). This analysis was performed on the reduced 6-metabolite model. The Shen et al. study developed their own model, which is plotted in black.(44)

A machine learning algorithm based on an ordinal response mixed effect model with LASSO regularization was used to generate our prediction model of the composite outcome measure using serum metabolic profiles and baseline patient demographics (age, sex and BMI) as combined inputs (Figure 5B). Proteins were not included in the model, since our study had around three times more metabolomics samples than proteomics samples. The final model included 32 metabolites and achieved an average concordance-index of d=0.69 (SE=0.017, 95% CI of [0.65, 0.72]), which is equivalent to a receiver operating characteristics area under the curve (ROC-AUC) on a binary outcome. This was a significant improvement (p-value = 3E-5) over a baseline model containing only age, sex, and BMI, which had a performance of d=0.59 (SD=0.02, 95%

CI of [0.56, 0.64]). The 32 metabolites in the final model included 14 amino acids, 6 nucleotides, 6 lipids and 2 metabolites related to energy metabolism (Figure 5C). Interestingly, our metabolite-based model showed improvement over the baseline model not only for predicting the composite outcome, but also for predicting some of its individual components (i.e., intubation, AKI, supplemental oxygen requirement, and length of hospital stay, Figure 5D).

We assessed the tradeoff between the number of metabolites incorporated into the model and the model’s prediction performance across a range of included metabolites. Our analysis revealed that most of the predictive power (d ∼ 0.68) was already achieved with the first six metabolites (cis-aconitic acid, hydrocinnamic acid, pantothenic acid, 7-methylguanosine, citrulline, methionine sulfoxide), after which prediction performance did not significantly improve (Supplementary Figure 4). This finding suggests that a targeted assay of six metabolites could predict disease severity with this accuracy, although independent validation in other datasets remains to be established.

As a validation step, we tested the performance of our reduced six-metabolite model on blood metabolomics data from two previously published studies by Su et al.(43) and Shen et al.(44) (Figure 5E). The Su study reported COVID-19 severity using a WHO-based score with 7 ordinal levels from mild to severe. Our model achieved a concordance index of d=0.83, again outperforming a model that just consisted of age, sex, and BMI (Figure 5E, left). The Shen study differentiated two groups of COVID-19 patients, mild and severe. That study also developed a prediction model, and we extracted their patient prediction scores from the paper to be able to calculate a concordance index for comparison. The models were tested on two test cohorts, called ‘C2’ and ‘C3’ from the original publication (Figure 5E, right). Our 6-metabolite model consistently outperformed both the score from Shen et al. as well as the baseline model with age, sex, and BMI. Notably, overall concordance scores in the Shen dataset were substantially higher than in our own dataset, both as reported in the original paper as well as in the validation of our own model on their data. We believe this effect occurs due to the complex nature of our composite outcome score as opposed to a simple yes vs. no classification of severity, the small sample size in Shen et al., and the reporting of an unvalidated training set AUC of 0.95 in their study. Overall, this analysis demonstrates that the model replicates in independent datasets with varying definitions of severity, and that our model provides slightly better results compared to the previously reported prediction model.

## Conclusion

The novel coronavirus has ravaged the global healthcare system due to its high transmissibility and unpredictable clinical course that often affects multiple organ systems. Moreover, the long-term consequences of COVID-19 infection remain poorly understood. A full understanding of the pathogenesis of COVID-19 will require an unraveling of the mechanisms of inflammation, immune dysfunction, endothelial cell injury and dysregulated coagulation that underlie this disease.

In our study, we used an integrative proteomic-metabolomic analysis to identify global molecular signatures specific to the acute illness of COVID-19, while many prior metabolomic and proteomic studies have not assessed the interplay between proteins and metabolites. Our analyses establish associations of specific inflammation and vascular injury-related proteins with various metabolites during COVID-19, which appear to link inflammation with mitochondria-dependent energy metabolism and viral replication, as well as coagulation with fibrogenesis and glycolysis.

Our discovered network modules not only provide a better understanding of disease pathogenesis, but also facilitate novel potential therapeutic targets for COVID-19. The modules identified various proteins and metabolites involved in inflammatory and vascular injury processes, such as MMP 12, Cathepsin D and RAGE which, to the best of our knowledge, have not yet been studied as targets for therapeutic intervention in COVID-19. Of note, our modules contained IL-6, which is already a mainstay of treatment for severe disease(50), and several other molecules such as carnitine, niacinamide and IFN-gamma which others have been studying in the context of COVID-19 therapies(51-53).

There is increasing evidence that evaluating symptoms and multiple clinical outcomes during acute disease is crucial in determining the risk of long COVID-19(54). To the best of our knowledge, we are the first group to develop a composite outcome measure in COVID-19 using multiple clinical indices in a prediction model that assesses not only COVID-19 disease severity but also the sequelae of COVID-19 that characterize post-acute COVID-19 syndrome (PACS). Compared to dichotomous outcome measures such as death and survival, our composite outcome score reflects a broader, more holistic assessment of COVID-19 morbidity in the hospital setting. Moreover, we were able to validate the model in two independent studies, thereby demonstrating its generalizability and translational potential.

Several key strengths underlie our study cohort. As opposed to the use of healthy controls reported in other COVID-19 studies(9, 20, 43, 44, 55), our use of non-COVID patient samples, in the same hospital during the same period between March and April 2020, allowed us to investigate the interactions highly relevant to COVID-19 pathogenesis and clinical course. Additionally, we analyzed a relatively larger cohort compared to other studies, with hundreds of samples available for both metabolomic and proteomic analysis.

Our study has several limitations. As alterations in proteome and metabolome were analyzed in sera but not in lung tissues or bronchoalveolar lavage fluid, our results may not reflect what occurs at tissue-specific cellular levels. Furthermore, based on the current study design and methodology, the correlative relationships we report between metabolomic, and proteomic alterations and SARS-CoV-2 outcomes should be interpreted as purely correlative rather than causal in nature. Additional studies are required to define the mechanistic roles of individual molecules highlighted in this paper. Finally, as our study was only a single center investigation, our results will need to be validated in other cohorts.

In conclusion, our investigation has sought to not only define the metabolomic and proteomic signatures of COVID-19, but also to explore interactions between metabolites and proteins that can serve as a roadmap for future mechanistic studies. We have furthermore proposed a novel clinical composite outcome score that can be used in a clinical prediction model for COVID-19. Ultimately, a better understanding of the pathophysiology of COVID-19 at the molecular level may lead to short-term and long-term targeted therapies.

## Methods

### Cohort description

This is a single-center prospective analysis of one cohort comparing hospitalized COVID-19 patients and non-COVID-19 controls. Our cohort was comprised of 330 patients with confirmed SARS-CoV-2 RT-PCR, and 97 non-COVID-19 controls with negative RT-PCR results who were hospitalized at the NewYork-Presbyterian Hospital/Weill Cornell Medical Center between March and April 2020. This study has been approved by the Weill Cornell Medicine (WCM) IRB with protocol #19-10020914. Remnant serum samples were matched with selected patients after which patients were deidentified. Controls were randomly selected patients admitted to the hospital with symptoms suspicious for COVID-19, but with negative SARS-CoV-2 RT-PCR. Seventy nine percent of control group patients had shortness of breath, fever, cough or chest pain which are commonly seen in COVID-19. Children (less than 18 years old) and pregnant women (confirmed by a positive beta-HCG test and/or medical records) were excluded.

### Sample handling

Standard practices for serum collection and storage at the NewYork-Presbyterian/Weill Cornell Medical College include collecting venous blood into a serum-separating tube (SST), and serum is obtained by centrifuging at 1,500g for 7 minutes as soon as possible with a maximum time limit of 2 hours from the time of collection. The specimens are typically stored at 4°C for 1 to 5 days before coded/de-identified and then transferred into a -80°C freezer. Samples were thawed and inactivated in different ways: for the metabolomic analysis, x3 sample volume of HPLC grade ethanol were added; for the proteomic analysis, the samples were heat-inactivated in a water bath of 56°C for 15 minutes. After these processes, the samples were stored again at -80°C until the analyses were performed.

### Data Collection

Data were obtained from the Weill Cornell Medicine COVID Institutional Data Repository (COVID-IDR), which is a high-quality registry of COVID-19 patients at NewYork-Presbyterian - Cornell with laboratory confirmed SARS-CoV-2 RT-PCR. The COVID-IDR houses both manually and automatically extracted Electronic Health Record (EHR) data. Demographics, comorbidities, and important dates of patients’ hospital course (admission, intubation, extubation, discharge, death) were extracted by a team of medical professionals and stored in the COVID-IDR. Laboratory tests, ventilation parameters, vital signs, and respiratory variables were additionally available via automated extraction through the Weill Cornell-Critical Care Database for Advanced Research (WC-CEDAR) within the COVID-IDR. WC-CEDAR(56) is a critical care database originally designed to automatically extract, transform, and store EHR data on Intensive Care Unit (ICU) patients; it was expanded to include all hospitalized patients during New York City’s COVID-19 surge. Data not available within WC-CEDAR were manually extracted and recorded in REDCap.

### Metabolomic profiling

Targeted Metabolite profiling was performed according to a method described in a previous publication (57). For metabolite extraction, 80 μL of pre-chilled methanol (−80 °C) was added to 20 μL of serum. The sample was vortexed for 1 min and then incubated at-80 °C for 2 hours before it was centrifuged at 20,000 g for 15 min at 4 °C to remove the pellet. The supernatant was transferred to a new Eppendorf tube and dried completely with a Speedvac for 30 min (with heat off). The dried sample was redissolved in HPLC grade water before it was applied to the hydrophilic interaction chromatography LC-MS. The sample injection order was randomized.

Metabolites were measured on a Q Exactive Orbitrap mass spectrometer (Thermo Scientific), which was coupled to a Vanquish UPLC system (Thermo Scientific) via an Ion Max ion source with a HESI II probe (Thermo Scientific). A Sequant ZIC-pHILIC column (2.1 mm i.d. × 150 mm, particle size of 5 μm, Millipore Sigma) was used for separation of metabolites. A 2.1 × 20 mm guard column with the same packing material was used for protection of the analytical column. Flow rate was set at 150 μL/min. Buffers consisted of 100% acetonitrile for mobile phase A, and 0.1% NH_4_OH/20 mM CH_3_COONH_4_ in water for mobile phase B. The chromatographic gradient ran from 85% to 30% A in 20 min followed by a wash with 30% A and re-equilibration at 85% A. The column temperature was set to 30 °C and the autosampler temperature was set to 4 °C. The Q Exactive was operated in full scan, polarity-switching mode with the following parameters: the spray voltage 3.0 kV, the heated capillary temperature 300 °C, the HESI probe temperature 350 °C, the sheath gas flow 40 units, the auxiliary gas flow 15 units. MS data acquisition was performed in the m/z range of 70–1,000, with 70,000 resolution (at 200 m/z). The AGC target was 3,000,000 and the maximum injection time was 100 ms. The MS data was processed using XCalibur 4.1 (Thermo Scientific) to extract the metabolite signal intensity for relative quantitation. Metabolites were identified using an in-house library established using chemical standards. Identification required exact mass (within 5ppm) and standard retention times. As a quality control, a mixture of standard compounds was injected thirteen times throughout the LC-MS data acquisition process for monitoring of the stability of LC retention time, the MS mass accuracy and the signal intensity. The median coefficient of variation for metabolite quantitation based on the quality control sample was 0.061. The data from the serum samples showed that both retention time and mass accuracy were highly stable throughout the experiment (Supplementary Figure 5).

### Proteomic profiling

Proteomics analysis was performed using the Olink platform (Uppsala, Sweden) at the Proteomics Core of Weill Cornell Medicine-Qatar, according to manufacturer’s instructions. We used the Inflammation, Cardiovascular II and Cardiovascular III panels. High throughput real-time PCR of reporter DNA lined to protein specific antibodies was performed on a 96-well integrated fluidic circuits chip (Fluidigm, San Francisco, CA). Each sample was spiked with quality controls to monitor the incubation, extension, and detection steps of the assay. Additionally, samples representing external, negative, and inter-plate controls were included in each analysis run. From raw data, real time PCR cycle threshold (Ct) values were extracted using Fluidigm reverse transcription polymerase chain reaction (RT-PCR) analysis software at a quality threshold of 0.5 and linear baseline correction. Ct values were further processed using the Olink NPX manager software (Olink, Uppsala, Sweden). Here, log2-transformed Ct values from each sample and analyte were normalized based on spiked-in extension controls and scale-inverted to obtain Normalized log2-scaled Protein Expression (NPX) values. NPX values were adjusted based on the median of inter plate controls (IPC) for each protein and intensity median scaled between all samples and plates.

Each metabolite and protein was annotated with pathways from the Kyoto Encyclopedia of Genes and Genomes (KEGG) database(59).

### Data preprocessing

Children, pregnant women, and samples after intubation were excluded from all analyses. The metabolomics data was measured in three different batches. For each batch, data was preprocessed by filtering out samples with more than 50% missing values, followed by filtering out metabolites with more than 25% missing values, probabilistic quotient normalization(60), and log2 transformation. Two extreme outlier metabolites were manually removed (phosphorylcholine and adenosine monophosphate). The next step was to merge the different batches into a joint dataset. Batch 3 contained only control patients and could thus not be simply added by batch correction. To avoid issues created by this imbalanced experimental design, batches 2 and 3 contained an overlapping set of samples which were used for an anchor-based normalization by dividing each metabolite in batch 3 by the mean fold change of the overlapping samples. The anchor samples from batch 3 were then deleted and the batches combined using median-based batch correction(61). Overall, this procedure eliminates batch effects and allows for a batch that only contains control samples. Missing values were then imputed using the k-nearest neighbor approach(61).

Proteomics data preprocessing included the same steps of filtering, quotient normalization, logging, and missing value imputation with identical parameters as for the metabolomics data. Ten proteins were measured as duplicates on the Olink platform, so their expression values were averaged.

### Statistical analysis

Differential expression of metabolites and proteins for both the COVID-19 vs. control analysis as well as the clinical parameter analysis within the COVID-19 cohort was assessed using the following linear mixed effect model:

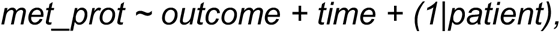

where *met_prot* is each individual metabolite or protein, *outcome* is either COVID-19 yes/no or the value of a clinical parameter, *time* is the day of sample taking as a factor, and *(1*|*patient)* is a random effect per patient to account for repeated measurements. P-values are reported for the significance of the outcome term.

Data preprocessing and statistical analysis was performed using the “maplet” toolbox for R(62) (https://github.com/krumsieklab/maplet).

### Network construction

The dataset was first reduced to the samples that overlap between metabolomics and proteomics (n=227), and corrected for age, sex, BMI, and COVID-19 status (yes/no). A Gaussian Graphical Model (GGM) based network was then constructed using the GeneNet algorithm(63) and drawing an edge for all partial correlations with an FDR smaller than 0.2. In a second step, this network was condensed to highlight the connections between molecules that were significantly different between COVID-19 and controls. To this end, a shortest-path distance matrix between all molecules was constructed and subset to the significant molecules. A minimum spanning tree(64) of this matrix was then constructed to visualize a simplified network.

### Composite Outcome

A detailed description of the construction of the composite outcome along with patient numbers in each group can be found in Supplementary Figure 3.

### Regularized linear mixed effect ordinal regression model

A new regression model was developed to deal with an ordinal outcome, repeated measurements, and feature selection with regularization. Repeated measurements are handled as a random effect, while age, sex, BMI, and metabolites are treated as fixed effects. The metabolites are penalized using an L_1_ LASSO-type regularization to obtain a sparse solution. The model is fitted using an mixed-effect ordinal regression model with complementary log-log link function(65), using maximum likelihood (ML) estimation as proposed by Ripatti and Pamgren(66). An optimal LASSO penalty parameter was estimated through an iterative algorithm for maximizing the Laplace approximation of the integrated model likelihood. This approach was adapted from Therneau(67), which was originally developed for mixed effect Cox models. To obtain an unbiased estimate of the model performance, leave-one-out-cross-validation across the entire dataset was performed. The added value of metabolomics data over baseline clinical data was assessed by comparing the final model with a model only consisting of age, sex, and BMI.

### Validation datasets

Metabolomics data were downloaded from Su et al.(43) (n=121) and Shen et al. (44) (containing two validation sets, n=10 and n=19). The Su dataset contained all six metabolites from our reduced model as well as age, sex and BMI as baseline parameters. The Shen study also covered age, sex and BMI, and the first test dataset (“C2”) contained all six metabolites. The second test dataset (“C3”) was measured using a targeted assay of only 7 metabolites and 22 proteins, and the metabolites did not overlap with our model metabolites. Thus, we had to follow a more complex procedure for validation. We first applied our risk score in their training cohort, which contained all metabolites. In the training cohort, this score was then regressed on the available measurements in C3, i.e. modeling ‘score ∼ metabolite1 + … + metabolite7 + protein1 + …+ protein22’. The coefficients from this model were then used in C3 to derive a surrogate severity score, which we evaluated in Figure 5.

## Supporting information

Supplementary Figure 1: Time Effect

Supplementary Figure 2: PCAs

Supplementary Figure 3: Composite Outcome

Supplementary Figure 4: Tradeoff

Supplementary Figure 5: Metabolomics Stability

Supplementary Table 3: Intubation-at-blood-draw Stats

Supplementary Table 4: Case-Control Stats

Supplementary Table 5: Clinicals Metabolomics Stats

Supplementary Table 6: Clinicals Proteomics Stats

Supplementary Data 1: Networks

## Data Availability

There are legal and ethical restrictions on data sharing because the Institutional Review Board of Weill Cornell Medicine did not approve public data deposition. The data set used for this study constitutes sensitive patient information extracted from the electronic health record. Accordingly, it is subject to federal legislation that limits our ability to disclose it to the public, even after it has been subjected to deidentification techniques. To request the access of the de-identified minimal dataset underlying these findings, interested and qualified researchers should contact Information Technologies & Services Department of Weill Cornell Medicine .

## Data Availability

The data used in this study can be downloaded at https://doi.org/10.6084/m9.figshare.19115972.v1.

## Code Availability

Code to reproduce all the statistical results presented in this paper is available at https://github.com/krumsieklab/covid-omics.

