## Supplementary Figure 1: Time Effect for "Integrative Metabolomic and Proteomic Signatures Define Clinical Outcomes in Severe COVID-19"

**Supplementary Figure 1 – Time effects across first three days of hospitalization**

For each metabolite and transcript, the same linear mixed effect model as for the COVID-19. vs normal analysis was computed: x~Time+Status+(1|Patient_ID). In this case we evaluated the significance the "Time" term to evaluate the effects of the day of sample taking.

### Metabolomics

A total of 35 out of 125 metabolites showed a significant time effect.

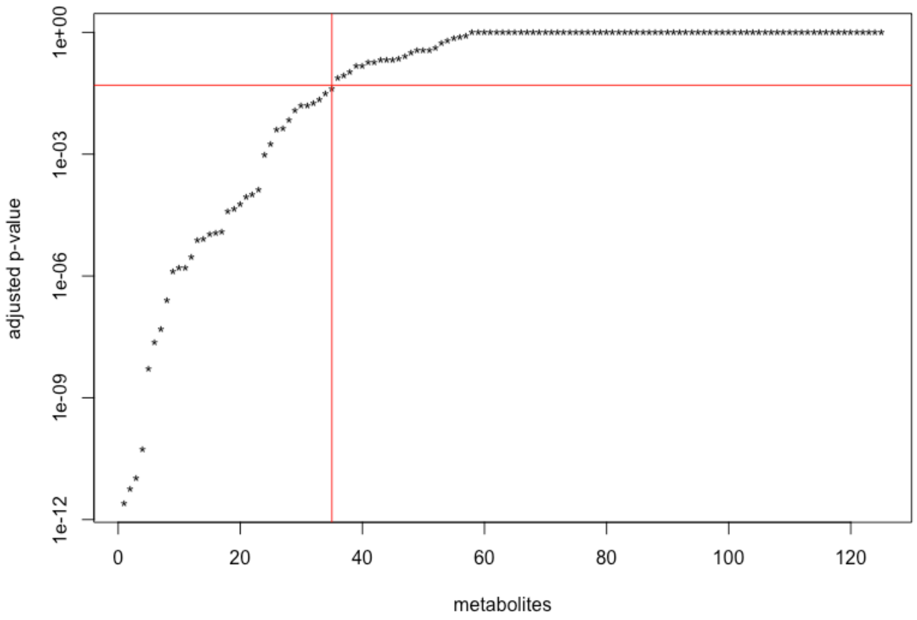


Examples, top 4 associations

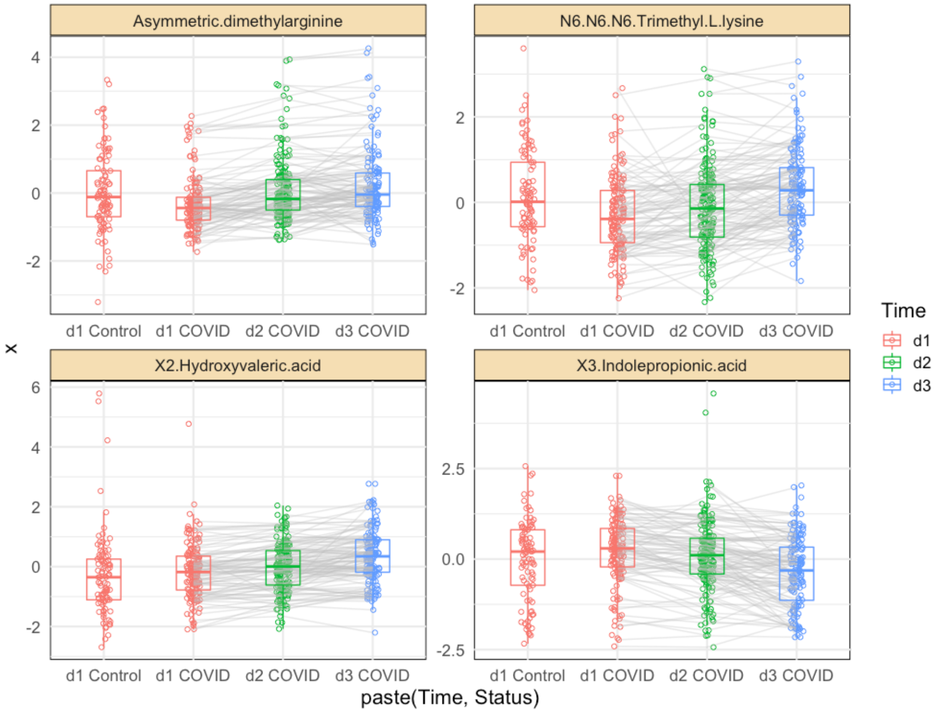

### **Proteomics**

A total of 25 out of 266 proteins showed a significant time effect.

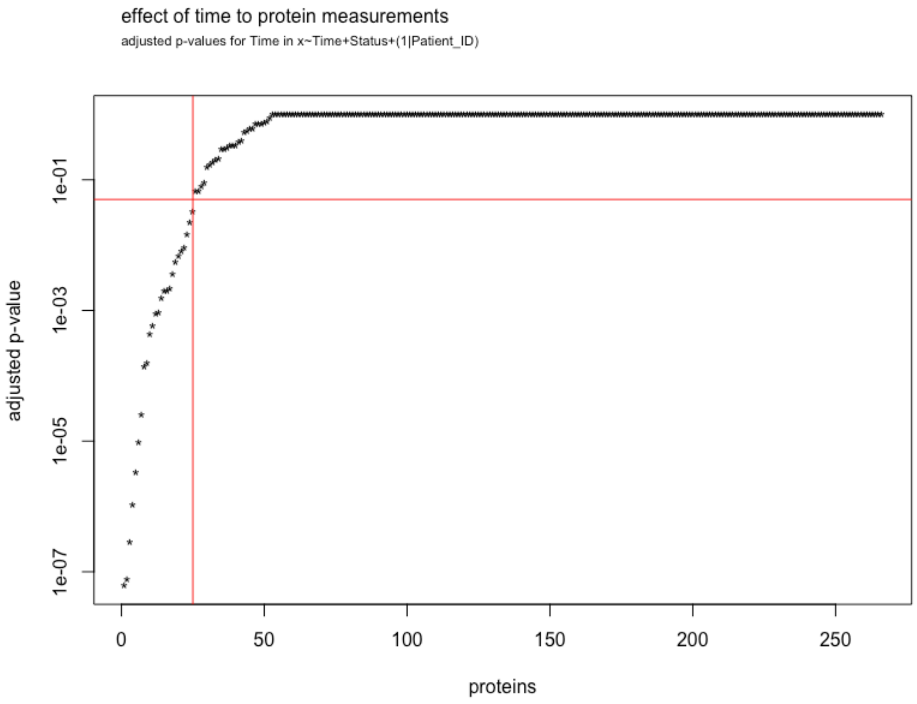


<u>Examples, top 4 associations</u>

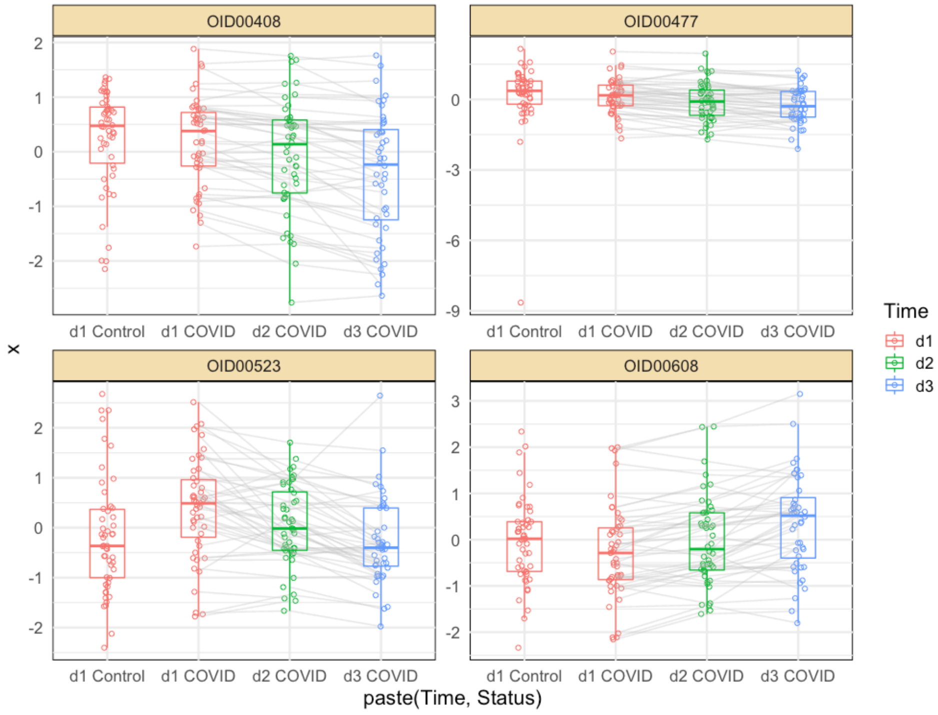
