## Supplementary Figure 2: PCAs for "Integrative Metabolomic and Proteomic Signatures Define Clinical Outcomes in Severe COVID-19"

### Supplementary Figure 2 – Sample-wise principal component analysis (PCA)

#### Metabolomics

Colored by status

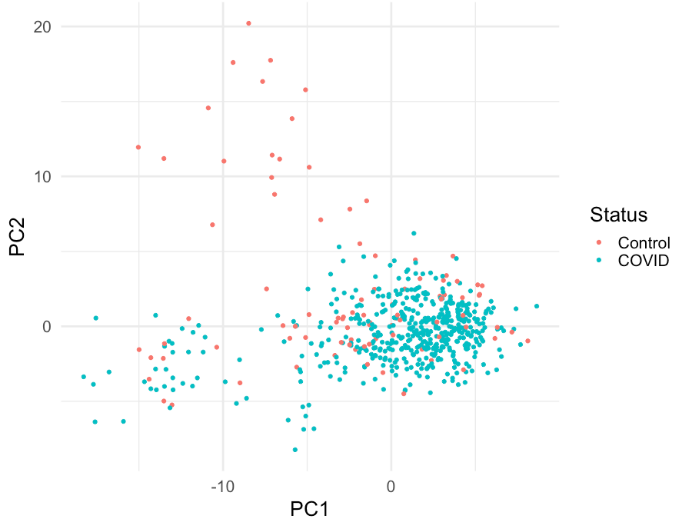


Colored by batch

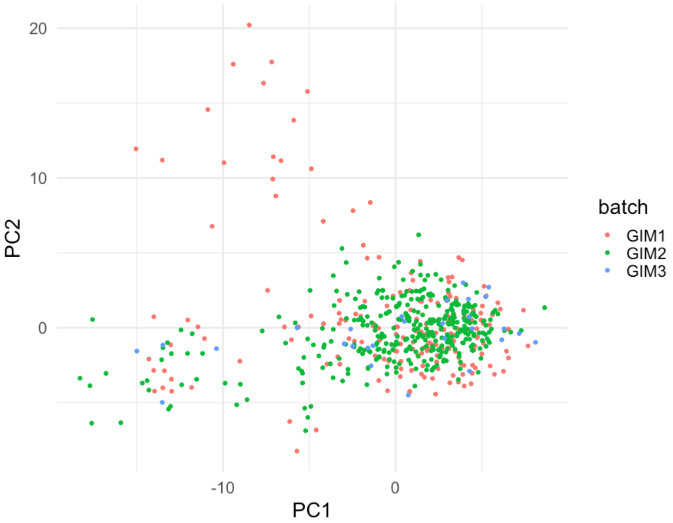


#### Proteomics

Colored by status

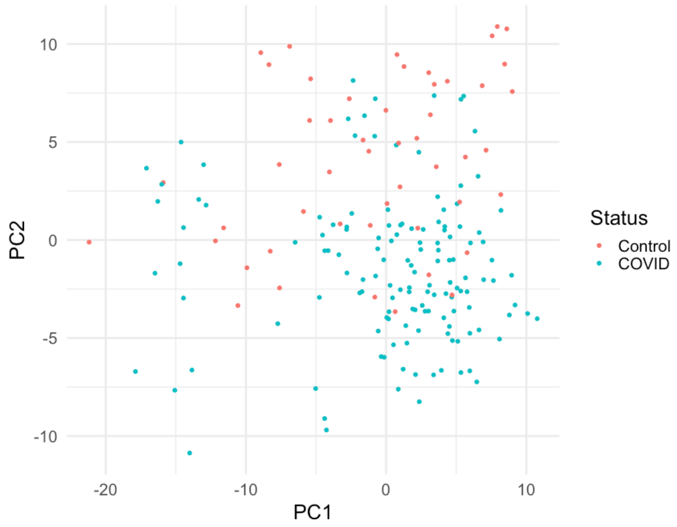


Colored by batch

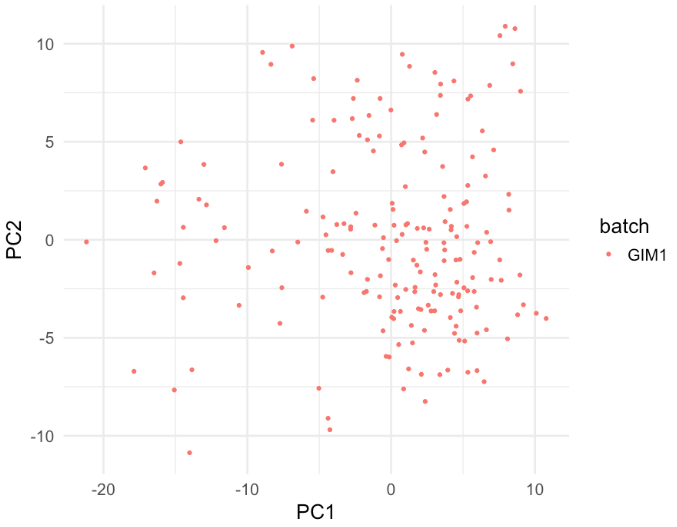
