## Supplementary Figure 3: Composite Outcome for "Integrative Metabolomic and Proteomic Signatures Define Clinical Outcomes in Severe COVID-19"

**Supplementary Figure 3 - Composite outcome definition**

The goal of the composite outcome is to order patients on an ordinal scale of disease severity. Death is the most severe event, followed by kidney replacement therapy (KRT) at discharge, prolonged organ failure support (POF support), and other clinical indices and events. To assign a severity score per patient, the list below is applied sequentially from top to bottom. For example, patients are first categorized into death yes and no. Within the survivor group, patients are ranked by kidney replacement therapy yes/no, then by prolonged organ failure yes/no, and so forth. We decided to include kidney injury (kidney replacement therapy and acute kidney injury) since kidney involvement in patients with COVID-19 is common and is associated with high mortality(1). Note that time-to-death is only used within the death-yes group. Further note that numeric indices do not fully resolve the ranking due to discrete numeric values.

1. Death (yes/no)
    a. within Death-yes group Time-to-death (days)
2. Kidney replacement therapy at discharge (yes/no)
3. Prolonged organ failure support (yes/no)*
4. Intubation (yes/no)
5. Kidney replacement therapy any (yes/no)
6. Oxygen Device (ordinal categories)**, ł
    i. MV: Mechanical Ventilation
    ii. NIMV: Non-invasive mechanical ventilation
    iii. HFNC: High-Flow nasal cannula
    iv. NR: Non-rebreather mask
    v. Venti-mask
    vi. NC or TC: Nasal cannula or trach collar
    vii. RA: Room air
7. FiO2 (2 decimal numeric value)***, ł
8. Acute Kidney Injury (ordinal categories as 0,1,2,3)
9. Hospitalization length (days)
10. Disposition (ordinal categories as Skilled Nursing Facility > Subacute rehab > Acute rehab > Home)

* Prolonged organ failure support is defined as Mechanical Ventilation > 14 days or Renal replacement therapy > 14 days.

** Oxygen Device and FiO2 within each device group defines Hypoxia.

*** FiO2 is used within 6 categories of O2 device

ł Oxygen Device and FiO2 are evaluated at the time of blood draw

The following figure illustrates the first ranking steps and the patient numbers associated with them.

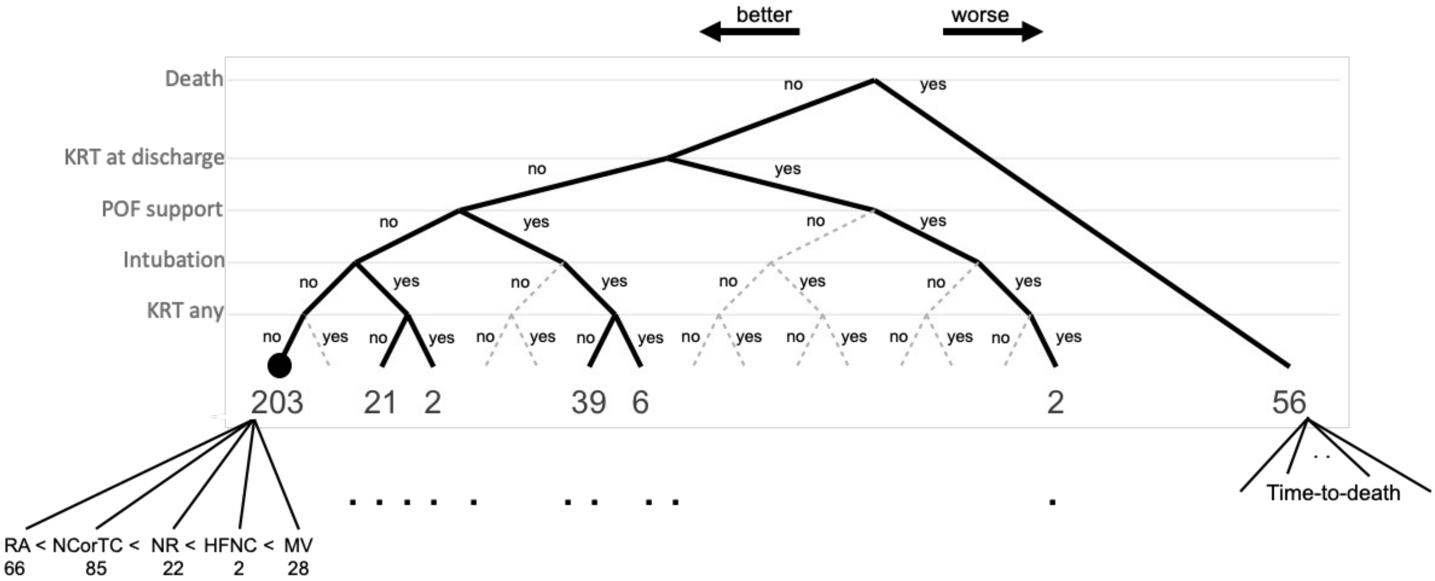


KRT = kidney replacement therapy. POF = prolonged organ failure support, RA = room air. NCorTC = Nasal cannula or trach collar. NR = non-rebreather mask. HFNC = high-flow nasal cannula. MV = mechanical ventilation
