## Supplementary Figure 4: Tradeoff for "Integrative Metabolomic and Proteomic Signatures Define Clinical Outcomes in Severe COVID-19"

### Supplementary Figure 4 – Tradeoff between metabolites included in model's performance

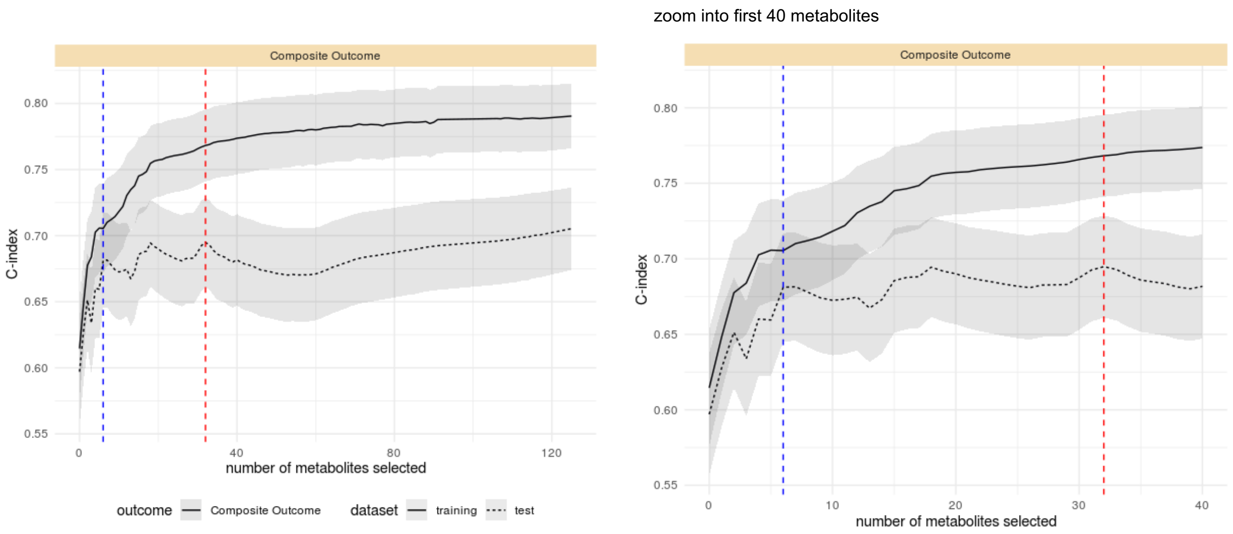


This figure shows the tradeoff between the selected number of metabolites and model performance. The optimal model had 33 metabolites (red line), while a sparser solution with similar performance can be achieved with 6 metabolites only (blue line).
