## Supplementary Figure 5: Metabolomics Stability for "Integrative Metabolomic and Proteomic Signatures Define Clinical Outcomes in Severe COVID-19"

**Supplementary Figure 5 – Assessment of reproducibility of the LC-MS metabolomics data.**

**A:** Coefficient of variation for quantitation calculated based on the quality control sample which was injected thirteen times throughout the serum sample runs. The median CV was 0.061. **B:** Overlaid LC retention time shift distributions of all the serum samples. For each sample, a density plot was generated using the retention time shifts of detected metabolites. **C+D:** Overlaid mass error distributions of all the serum samples. For each sample, a density plot was generated using the mass errors of detected metabolites. The plot was generated separately for positive and negative ion modes.

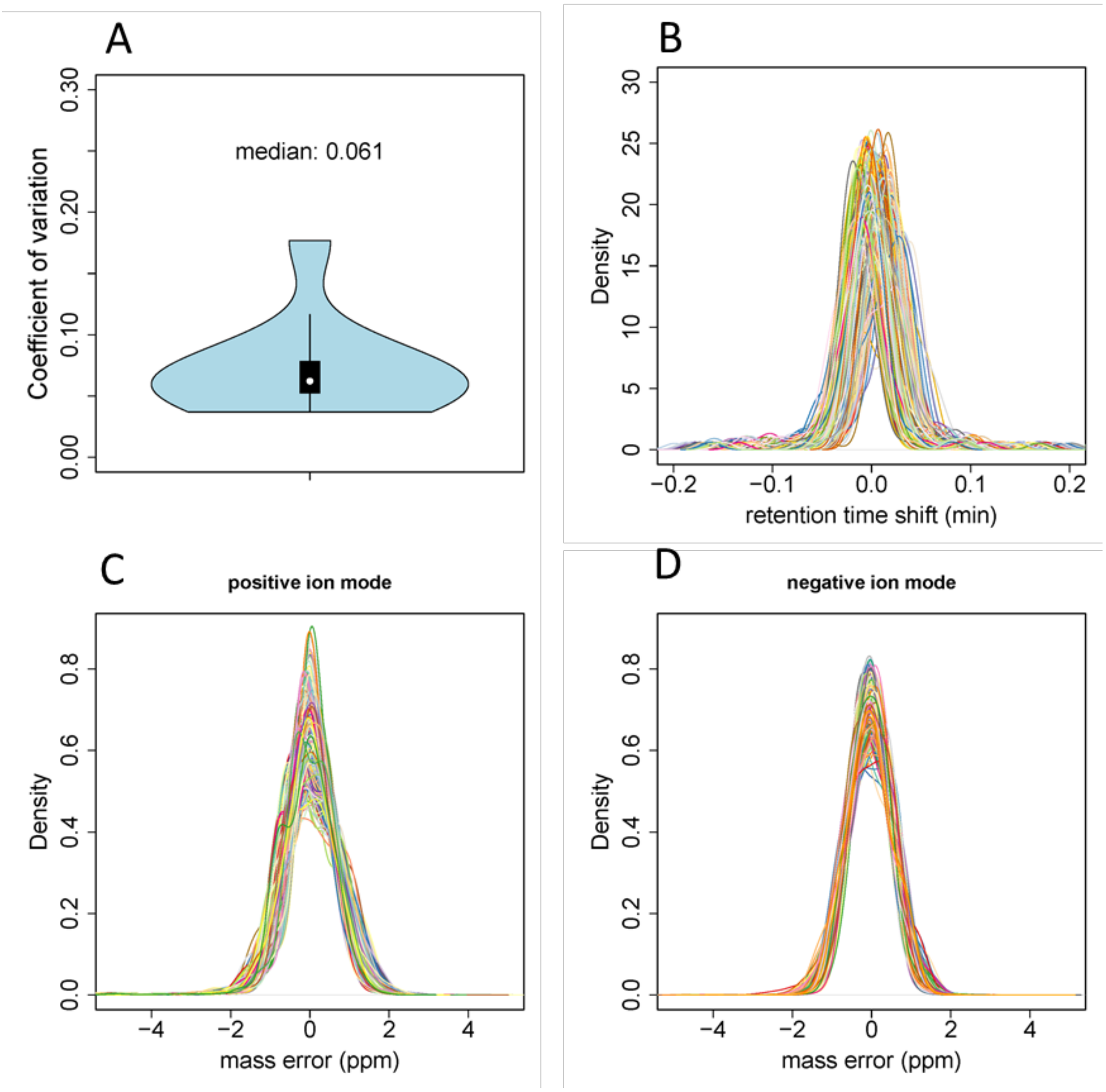
