## Supplementary Table 1 for "Integrative Metabolomic and Proteomic Signatures Define Clinical Outcomes in Severe COVID-19"

### Supplementary Table 1: Additional clinical characteristics of the cohort

| Characteristic | Negative, N = 97[1] | COVID-19, N = 330[1] | p-value[2] |
|---|---|---|---|
| **Laboratories** | | | |
| White blood cell | 7.6 (4.9, 11.6) | 6.3 (4.6, 8.6) | **0.015** |
| Neutrophils | 4.9 (2.9, 9.0) | 4.8 (3.2, 7.0) | 0.8 |
| Hemoglobin | 10.80 (9.00, 13.00) | 13.00 (11.80, 14.20) | **<0.001** |
| Neutrophil/Lymphocytes | 5 (3, 9) | 6 (4, 10) | 0.2 |
| Partial thromboplastin time | 30.2 (27.9, 33.8) | 31.8 (29.4, 34.1) | 0.11 |
| Prothrombin time | 13.35 (11.85, 15.80) | 13.45 (12.60, 14.47) | 0.5 |
| Fibrinogen | 434 (369, 454) | 461 (394, 628) | 0.4 |
| Albumin | 3.10 (2.50, 3.70) | 3.00 (2.60, 3.40) | 0.14 |
| Total bilirubin | 0.50 (0.30, 0.85) | 0.60 (0.40, 0.80) | 0.4 |
| Lactate dehydrogenase | 328 (218, 480) | 408 (308, 521) | 0.092 |
| Creatinine | 1.01 (0.75, 1.38) | 0.94 (0.76, 1.23) | 0.5 |
| Lactate | 1.30 (1.00, 1.87) | 1.17 (0.88, 1.65) | 0.2 |
| Glucose | 111 (93, 144) | 111 (95, 146) | 0.6 |
| Erythrocyte sedimentation rate | 46 (30, 60) | 63 (40, 86) | 0.1 |
| PH | 7.45 (7.45, 7.51) | 7.38 (7.32, 7.43) | **0.003** |
| PaO2 | 112 (86, 152) | 39 (25, 70) | **<0.001** |
| PaCO2 | 36 (30, 42) | 40 (35, 46) | 0.2 |
| **Clinical outcomes** | | | |
| Highest level of supplemental oxygen in the first 3 hours of admission | | | **0.001** |
| HFNC/NRB/NIV | 6 (6.2%) | 38 (12%) | |
| Mechanical ventilation | 3 (3.1%) | 15 (4.5%) | |
| Nasal Cannula | 16 (16%) | 107 (32%) | |
| None | 72 (74%) | 170 (52%) | |
| CXR results at admission | | | **<0.001** |
| Bilateral infiltrates | 14 (14%) | 232 (70%) | |
| Clear | 47 (48%) | 36 (11%) | |
| Not specified | 6 (6.2%) | 0 (0%) | |
| Other | 11 (11%) | 56 (17%) | |
| Pleural effusion | 0 (0%) | 6 (1.8%) | |
| Unilateral infiltrates | 19 (20%) | 0 (0%) | |
| ARDS diagnosis | 0 (0%) | 111 (34%) | **<0.001** |
| Respiratory co-infection | 41 (42%) | 57 (17%) | **<0.001** |

[1] Statistics presented: median (IQR); n (%)

[2] Statistical tests performed: Wilcoxon rank-sum test; Fisher's exact test; Fisher's test with simulated p-value; chi-square test of independence
