## Supplementary Table 2 for "Integrative Metabolomic and Proteomic Signatures Define Clinical Outcomes in Severe COVID-19"

### Supplementary Table 2: Symptoms and admission diagnosis of controls

| Characteristic | Negative, N = 97[1] |
|---|---|
| **Main symptom** | |
| Shortness of breath | 30 (31%) |
| Fever | 28 (29%) |
| Cough | 15 (15%) |
| Chest pain | 4 (4.1%) |
| Abdominal pain | 3 (3.1%) |
| Altered mental status | 3 (3.1%) |
| Dizziness | 3 (3.1%) |
| Arthralgia | 1 (1.0%) |
| Asymptomatic | 1 (1.0%) |
| Burn | 1 (1.0%) |
| Fall | 1 (1.0%) |
| Fatigue | 1 (1.0%) |
| Leg pain | 1 (1.0%) |
| Myalgias | 1 (1.0%) |
| Rhinorrhea | 1 (1.0%) |
| Seizure | 1 (1.0%) |
| Syncope | 1 (1.0%) |
| Tachycardia | 1 (1.0%) |
| **Diagnosis** | |
| Pneumonia | 27 (28%) |
| Acute coronary syndrome | 5 (5.2%) |
| Bacteremia | 5 (5.2%) |
| Heart failure exacerbation | 5 (5.2%) |
| Urinary tract infection | 4 (4.1%) |
| Asthma | 3 (3.1%) |
| Leukemia/Lymphoma | 3 (3.1%) |
| Upper respiratory infection | 3 (3.1%) |
| Multiple myeloma/Myelodysplastic syndrome | 2 (2.1%) |
| Neutropenic fever | 2 (2.1%) |
| Pulmonary edema | 2 (2.1%) |
| Pulmonary embolism | 2 (2.1%) |
| Serositis | 2 (2.1%) |
| Viral gastroenteritis | 2 (2.1%) |
| Abnormal vaginal bleeding | 1 (1.0%) |
| Acute cocaine intoxication | 1 (1.0%) |
| Acute limb ischemia | 1 (1.0%) |
| Acute psychotic episode | 1 (1.0%) |
| Alcohol withdrawal | 1 (1.0%) |
| Anxiety | 1 (1.0%) |
| Aortic aneurysm | 1 (1.0%) |
| Atrioventricular block | 1 (1.0%) |
| Bowel obstruction | 1 (1.0%) |
| Burn | 1 (1.0%) |
| Cellulitis | 1 (1.0%) |
| Cholecystitis | 1 (1.0%) |
| COPD exacerbation | 1 (1.0%) |
| Deep venous thrombosis | 1 (1.0%) |
| Dehydration | 1 (1.0%) |
| Diabetic ketoacidosis | 1 (1.0%) |
| Failure to thrive | 1 (1.0%) |
| Fever of unknown origin | 1 (1.0%) |
| Fracture | 1 (1.0%) |
| Guillain-Barré syndrome | 1 (1.0%) |
| Intracranial hemorrhage | 1 (1.0%) |

1n (%)
